# Psychophysiologic symptom relief therapy (PSRT) for post-acute sequelae of COVID-19: a non-randomized interventional study

**DOI:** 10.1101/2022.10.07.22280732

**Authors:** Michael Donnino, Patricia Howard, Shivani Mehta, Jeremy Silverman, Maria J Cabrera, Jolin B Yamin, Lakshman Balaji, Rebecca Tolin, Katherine M Berg, Robert Edwards, Anne V Grossestreuer

## Abstract

**Objective:** To determine if psychophysiologic symptom relief therapy (PSRT) will reduce symptom burden in patients suffering from post-acute sequelae of COVID-19 (PASC) who had mild/moderate acute COVID-19 disease without objective evidence of organ injury.

**Patients and Methods:** Twenty-three adults under the age of 60 with PASC for at least 12 weeks following COVID-19 infection were enrolled in an interventional cohort study conducted via virtual platform between May 18, 2021 and August 7, 2022. Participants received PSRT during a 13 week (approximately 44 hour) course. Participants were administered validated questionnaires at baseline and at 4, 8, and 13 weeks. The primary outcome was change in somatic symptoms from baseline, measured using the Somatic Symptom Scale-8 (SSS-8).

**Results:** The median duration of symptoms prior to joining the study was 267 days (IQR: 144, 460). The mean SSS-8 score of the cohort decreased from baseline by 8.5 (95% CI: 5.7-11.4), 9.4 (95% CI: 6.9-11.9), and 10.9 (95% CI: 8.3-13.5) at 4, 8, and 13 weeks respectively (all p<0.001). Participants also experienced statistically significant improvements across secondary outcomes including changes in dyspnea, fatigue, and pain (all p<0.001).

**Conclusion:** PSRT may effectively decrease symptom burden in patients suffering from PASC without evidence of organ injury. The study was registered on clinicaltrials.gov (NCT 04854772).

## Introduction

As of August 12, 2022, SARS-CoV-2 has infected over 91 million people in the United States, with a significant number of people experiencing prolonged symptomatology after their initial acute infection (1). The World Health Organization has defined post-acute sequelae of COVID-19 (PASC) as occurring in individuals with a history of probable or confirmed COVID-19, usually three months from onset of infection with symptoms lasting for ≥2 months and not explained by alternative diagnoses (2). According to various reports, 2-4 million Americans have been unable to work due to these symptoms (3), with $2.6 trillion projected in costs to the economy (4). Chronic pain in one or more body parts and chronic fatigue are some of the most frequently reported PASC symptoms (5–7). While reports differ depending upon definitions and methodology, previous studies of PASC estimate an incidence rate ranging from 4-35% of those diagnosed with COVID-19 (8–10). PASC can fall into several categories, with one being defined by organ injury typically after severe, acute COVID-19 disease. Another category of PASC consists of mild or moderate acute infection with chronic symptomatology but no clearly identifiable organ injury (i.e., normal chest imaging, normal pulmonary function tests, normal cardiac echocardiology, etc.) (6).

The etiology of symptoms for patients with PASC who had mild/moderate disease without evidence of organ injury by traditional clinical testing remains elusive. There have been a number of reports on the potential etiologies of PASC including capillary microclots (11), viral reservoirs in the gut (12), low cortisol levels (13), mitochondrial dysfunction (14), endothelial dysfunction (15), subtle changes on cardiac MRI (16), autoimmune reactions (6), and presence of a persistent virus/spike protein (17). In contrast to these reports, Sneller et al. examined 186 adults at least 6 weeks after laboratory-confirmed COVID-19 infection and found no evidence of persistent viral infection, autoimmune reaction, or abnormal immune activation (6). They did note an association of anxiety with the development of PASC similar to a recent report by Wang et al. (18). However, the concept that anxiety may be a driving factor thus implying that patients are in control of symptomatology may be dismissive and fails to explain some reported physiologic changes such as tachycardia or postural orthostatic hypotension (POTS). One explanation that incorporates both a physical and psychological conceptual model is a psychophysiologic response or syndrome. Psychophysiologic responses or syndromes are those which have physical/physiological outputs yet are centrally mediated. To date, most psychophysiologic processes have been best-described and accepted in the acute setting with the physiologic outputs ranging from benign (e.g., blushing response) to mildly pathologic (e.g., vasovagal syncope) to potentially life-threatening (e.g., Takotsubo’s cardiomyopathy). Chronic psychophysiologic processes tend to be less recognized but have been recently described in some chronic pain syndromes (19,20) and can be seen in entities such as Post-Traumatic Stress Disorder (PTSD) (21–23).

We previously proposed and tested the hypothesis that some forms of chronic pain in which a specific physiological etiology for the pain is unclear, such as chronic back pain, represent a psychophysiological process (19,20). Our scientific premise, based on initial encounters with PASC patients without organ injury and an assessment of the literature (see discussion), is that patients with mild-moderate initial COVID-19 infections without identifiable organ injury could potentially be suffering from a psychophysiologic process developed during or shortly after acute infection (15–18). Our psychophysiologic intervention in back pain patients significantly reduced back pain across multiple metrics; therefore, we hypothesized that a modified version of this intervention (Psychophysiologic Symptom Relief Therapy, PSRT) would attenuate symptoms in PASC patients across multiple health symptom domains. To test our hypothesis, we evaluated the efficacy of PSRT in reducing somatic symptom burden as measured by Somatic Symptom Scale-8 (SSS-8). We secondarily examined the impact of PSRT on chronic fatigue, functional activity, dyspnea, average pain, pain intensity, anxiety from pain, pain interference with life activities, gastrointestinal symptoms, and “brain fog.”

## Methods

### Study Design and Setting

This was an interventional cohort study. All participants received PSRT and served as their own control. Participant medication regimens were maintained by the participant and their medical team without interference by the study team and no pain or other medication prescriptions were prescribed or administered by the research team during the study.

This study was conducted remotely via video on a HIPAA-compliant virtual platform approved by the Institutional Review Board of Beth Israel Deaconess Medical Center in Boston, Massachusetts, from May 2021 to August 2022. All participants provided written informed consent. The study was registered on clinicaltrials.gov (NCT 04854772). Participants were recruited through physician referrals, flyers, and social media.

### Inclusion Criteria

Adults aged 18 to 60 years old with new symptoms attributed to PASC, such as extremity pain, dyspnea, headaches, chest pain, and fatigue occurring after an acute phase of COVID-19 disease, confirmed by a positive SARS-CoV-2 antigen or PCR test (or confirmed SARS-CoV-2 antibodies prior to vaccination), were included. PASC symptoms must have persisted ≥12 weeks after the end of the acute COVID-19 infection and must have persisted *≥*1 month without identified organ damage or an identified organic disease unrelated to COVID-19. Eligible symptomatology was defined as scoring ≥3 on the Somatic Symptom Scale-8 with symptoms persisting for ≥4 days/week. Participants were assessed for willingness to consider a mind-body intervention during a screening interview; only those who consented to this type of intervention were enrolled.

### Exclusion Criteria

Participants were excluded if they were >60 years of age (due to increased risk of organic symptom etiology) or had diagnosed non-COVID-19 organic disease as cause of pain, such as (but not limited to) malignancy, neurologic disorders (e.g., amyotrophic lateral sclerosis), or autoimmune disease. Patients with previous severe COVID-19 disease, defined as having been admitted to the ICU or by objective evidence of ongoing organ injury (e.g., persistent chest radiographic abnormalities, myocarditis), were excluded. Participants suffering from chest pain or dyspnea with identified lung or cardiac injury (e.g., chest radiograph abnormalities, cardiac ultrasound showing myocarditis, depressed ejection fraction) were excluded. Patients diagnosed with significant psychiatric comorbidities (e.g. schizophrenia, dementia) were also excluded.

### Intervention

PSRT is based on a psychophysiologic approach to understanding and treating pain, driven by the hypothesis that nonspecific pain can be the somatic manifestation of psychophysiological processes exacerbated by stress, repressed emotions, and other psychological processes (24,25). We recently published a randomized controlled study that showed PSRT led to substantial, statistically significant reduction in pain bothersomeness and pain-related disability in people suffering from nonspecific chronic back pain (19). We applied this treatment paradigm to the nonspecific symptoms common to PASC, including pain, in this trial. The PSRT course was led by a physician (author M.W.D.) and a mind-body expert (author P.H.).

The goal of PSRT is to address underlying stressors and psychological contributors (such as underlying conflicts and aversive affective states) to nonspecific symptoms in order to mitigate conditioned symptom responses and fear-avoidant behaviors. Weeks zero through four of the course included group classes twice per week (90-120 minutes per class) and consisted of psychophysiologic education, desensitization (including visualization techniques), and emotional awareness exercises (such as expressive writing). Education focused on providing participants with information about the role of stress and psychological processes in precipitating and perpetuating physical symptoms. Using desensitization exercises, participants were encouraged to approach, rather than avoid, physical activities first through visualizing an action that typically induces symptoms and then through physical exposures of feared symptom-inducing activities.

The final 9 weeks of PSRT is the Mindfulness Based Stress Reduction (MBSR) protocol as outlined by the Center for Mindfulness at the University of Massachusetts (26). This portion consisted of classes once per week for a duration of 90 to 120 minutes and focused on providing participants with mindfulness skills such as practicing awareness of breath, body scan, and sitting meditation. No new elements of PSRT were introduced but reminders about elements of the work were provided during the MBSR course. The detailed protocol of PSRT is described in the Supplementary Materials.

## Sample Size Justification

Due to the novelty of PASC, data on our primary outcome, the Somatic Symptom Scale (SSS-8), was not available in this population. We therefore used the overall population mean SSS-8 score (3.2±4.0, (27)) as the value that we hypothesized would be meaningful for participants to return to, and an estimate of the return to that population mean as representing a 45% reduction from baseline, based on a prior study in back pain (19). Assuming a baseline SSS-8 of 5.8±4.0 and a 13 weeks SSS-8 of 3.2±4.0, 22 participants provided 83% power with α=0.05 to detect this difference.

## Statistical Analysis

Descriptive statistics used frequencies and percentages for categorical variables and either means with standard deviations or medians with interquartile ranges (IQR) for continuous data, based on distribution. To compare differences within participants over time, a paired Student’s t-test or Wilcoxon signed-rank test was performed for continuous data, as appropriate. No adjustment was made for multiple comparisons. Statistical analyses were performed using R Statistical Software (v4.1.1; R Core Team 2021), Stata 17.0 (College Station, TX), and a two-sided p-value <0.05 was considered significant.

## Measures

Electronic surveys delivered through RedCap were administered at baseline (0 weeks) and subsequently at 4, 8, and 13 weeks post-enrollment. All of the scales described below were administered at each time point.

### Primary Outcome

Change in somatic symptoms was assessed by the Somatic Symptom Scale (SSS-8) (27). The SSS-8 is an eight item scale in which participants rank how bothered they have been by eight separate groups of symptoms over the past week on a scale of 0-4 from “not at all” to “very much” (19). Results from the SSS-8 are summed to an overall score that ranges from 0 to 32. Scoring of the SSS-8 is categorized as follows: no to minimal (0-3 points), low (4-7 points), medium (8-11 points), high (12-15 points), and very high (16-32 points) somatic symptom burden (27).

### Secondary Outcomes

Secondary outcomes included changes in average pain, pain intensity, anxiety from pain, fatigue, dyspnea, gastrointestinal (GI) distress (as measured in the SSS-8), “brain fog” and physical functioning. These changes were measured by the Brief Pain Inventory Questionnaire (BPI) (28– 30), Pain Anxiety Symptom Scale (PASS-20) (31), Fatigue Severity Scale (FSS-9) (32), Multidimensional Dyspnea Profile (MDP) (33), the GI component of the SSS-8, and Patient-reported Outcomes Measurement Information System (PROMIS-29) (34).

“Brain fog” was measured by an additional question in the traditional SSS-8 format asked at every time point of “During the past 7 days, how much have you been bothered by brain fog (difficulty thinking or concentrating)?” with the options of not at all, a little bit, somewhat, quite a bit, and very much. Of note, this addition is not formally part of the SSS-8 and has not been validated so we report this as a separate entity from the summation SSS-8 score.

The Brief Pain Inventory Questionnaire (Short Form; BPI) is a 9 item instrument that measures pain intensity and interference in daily life (28–30). Participants were asked to highlight areas where they experience pain on a full body diagram and respond to nine questions regarding pain intensity in which participants ranked their pain from 0 (“No Pain”) to 10 (“pain as bad as you can imagine”). This was followed by a seven item assessment of pain interference in which participants ranked how their pain has interfered with facets of daily life (e.g. General Activity, Mood, etc.) on a scale of 0 (“Does not Interfere”) to 10 (“Completely Interferes”). Pain intensity was calculated by averaging the scores from the first four elements (average pain, worst pain, least pain, and current pain) of the BPI. Pain interference was calculated by summing the seven rated components.

The Pain Anxiety Symptom Scale (PASS-20) is a 20 item questionnaire which measures a participant’s anxiety and fear as it relates to their pain (31). Participants were asked how often they engage in thoughts or activities (e.g. “I think that if my pain gets too severe, it will never decrease”). Each item was ranked on a six point frequency scale from 0 (“Never”) to 5 (“Always”). The scores from each question were summed to create an overall score and range from 0 to 100.

The Fatigue Severity Scale (FSS-9) (32) is a nine item scale that includes a visual analogue fatigue scale in which participants ranked their “global fatigue” on a sliding scale from 0 (“worst”) to 10 (“normal”). In the FSS-9, participants ranked how statements regarding fatigue (e.g., “Fatigue interferes with my physical functioning”) relate to their life in the past week on a scale of 1 (Strong Disagree) to 7 (Strongly Agree). The FSS-9 was calculated by summing all of the responses except those for the first question. Scoring ranges from 0 to 63.

The Multidimensional Dyspnea Profile (MDP) is a four item scale that assesses multiple qualities surrounding breathing including general discomfort and emotional response (33). Throughout this assessment, participants were asked to consider the last 7 days and answer each question focusing on that time frame. Participants were first asked to rank the general discomfort of breathing on a scale of 0 (“neutral”) to 10 (“unbearable”). They then ranked the intensity of their breathing sensations and their emotional experience during dyspneic episodes on a scale of 0 (“None”) to 10 (“As Intense as I can imagine”/”The most I can imagine”). The overall score was summed and ranges from 0 to 110.

We used the physical subsection of the Patient-Reported Outcomes Measurement Information System (PROMIS-29 v2.1) (34,35), a four item questionnaire in which participants are asked about their physical functionality. Participants ranked their physical function ability (e.g.,. “Are you able to do chores such as vacuuming or yard work”) on a four point scale ranging from “without any difficulty” (assumed 0) to “with much difficulty” (assumed 4). These scores were summed to represent an overall score, ranging from 0 to 16.

For attendance, percent attendance was calculated for each participant using the number of classes attended divided by the number of classes offered. If a participant joined the class for at least half of the duration, they were marked as having attended the class.

One of the techniques used during PSRT includes visualization of activities that typically trigger symptoms (visual motor imagery). During the visualization test, participants were asked to remain stationary and imagine an activity or movement that tends to result in symptoms. We evaluated the percentage of participants who were able to reproduce their symptoms through visualization.

## Results

### Participant Flow Through the Study and Demographics

Two hundred and six participants were screened for enrollment. Of these, 183 did not meet inclusion criteria or met exclusion criteria and 23 were enrolled (Figure 1). All participants completed the protocol except for one who discontinued attending classes yet completed surveys through 13 weeks between May 18, 2021 and August 7, 2022. The mean participant age was 42.6±9.6 years, 14 (61%) were female, and the median duration of PASC symptoms was 267 (IQR: 144, 460) days. Baseline characteristics are displayed in Table 1. All participants completed surveys at all timepoints and had complete capture of the outcomes unless noted below. The median percent attendance of classes was 100% (IQR: 90%, 100%), with 16 (70%) of participants attending all classes; the range for those who did not attend all classes was 50-95% attendance (mean of 78%±17% in those without perfect attendance). Seven groups of classes were held, ranging in size from one subject to seven subjects, with a median group size of 3 (IQR: 2, 5). Prior to entering the study, participants had tried a large variety of different therapies (Supplemental Table 1).

**Table 1.**
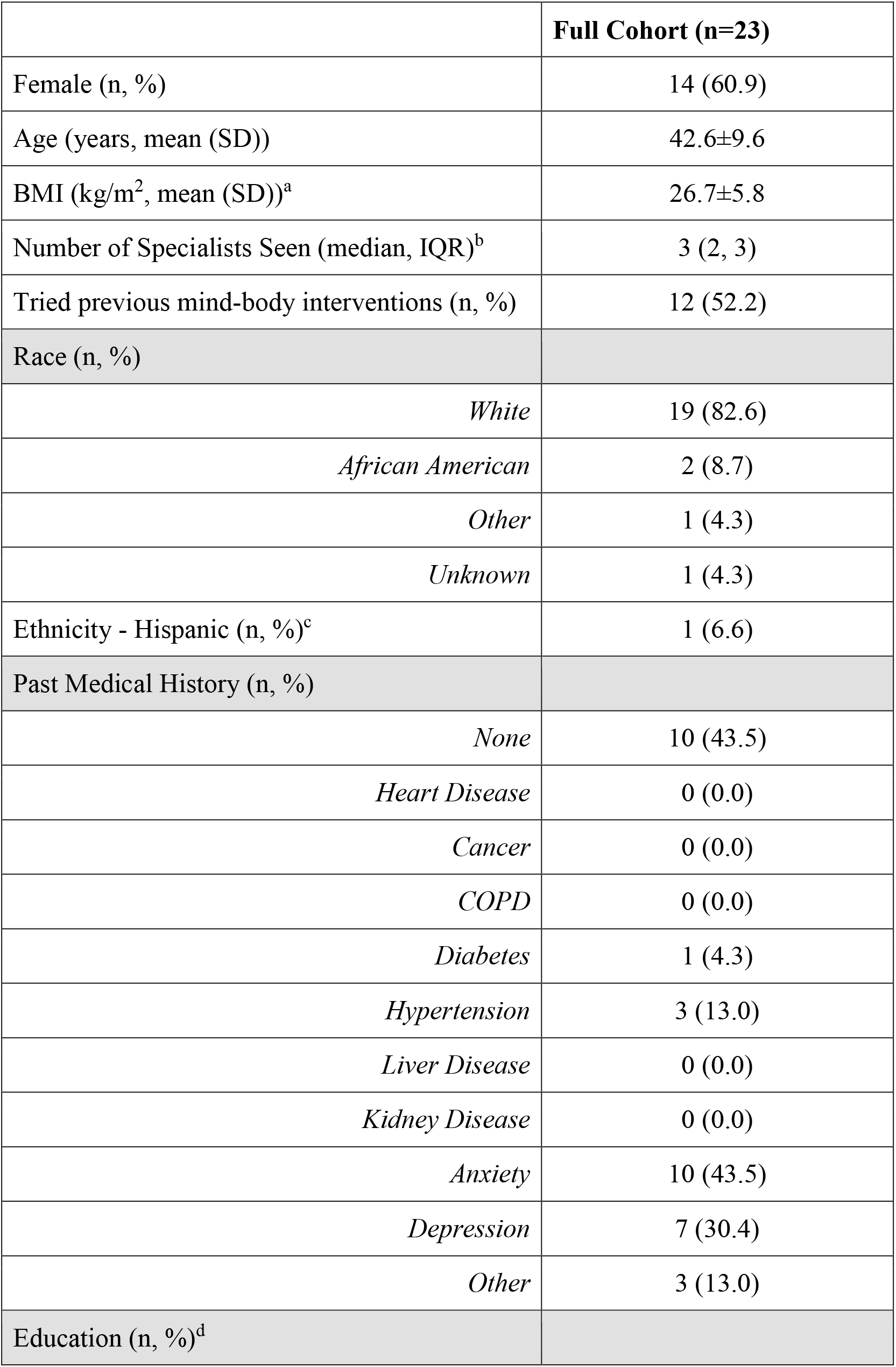
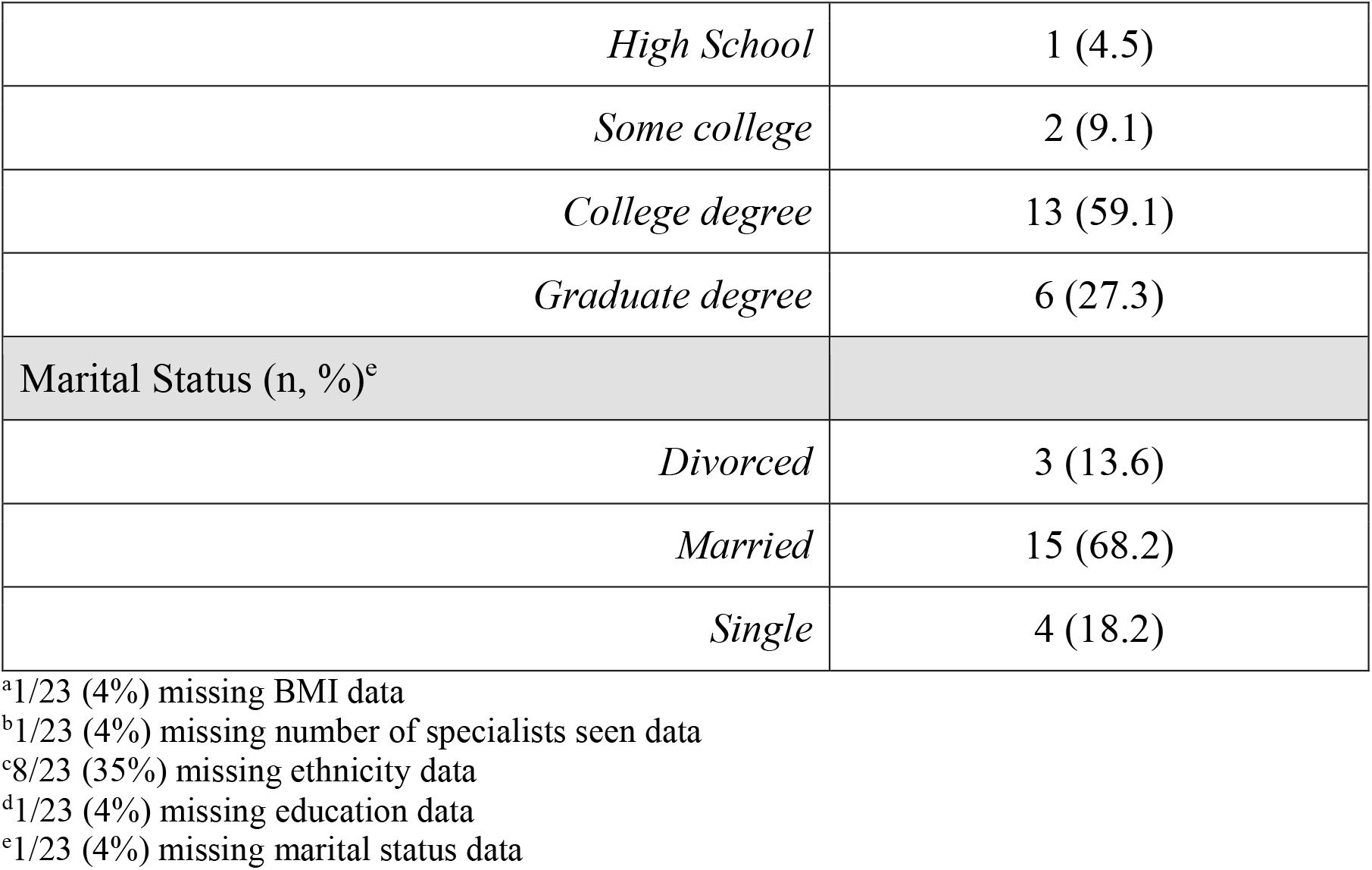
Participant demographics.

**Figure 1.**
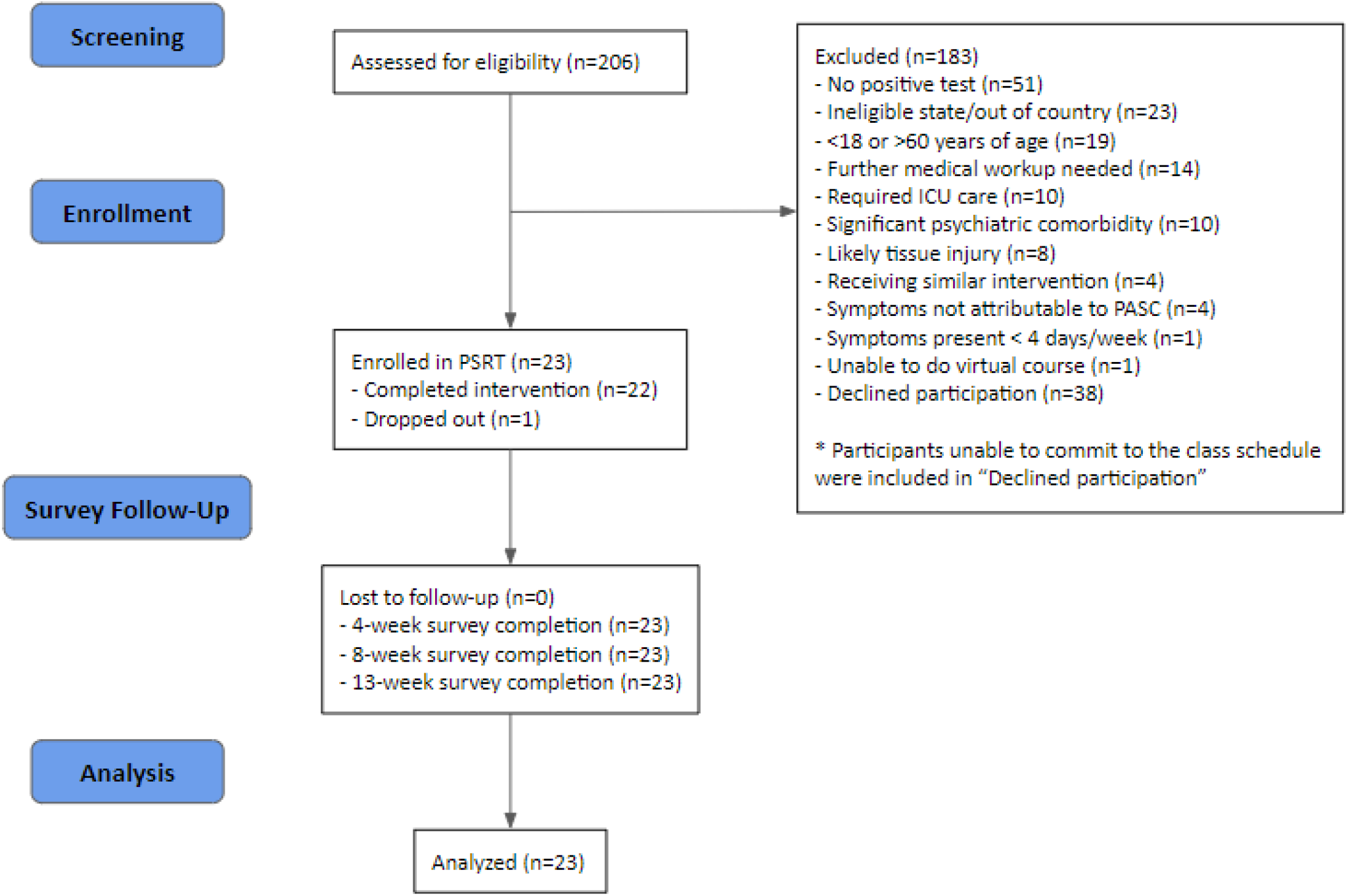
Consort Diagram.

### Primary Outcome

For the primary outcome of SSS-8, we found a statistically significant reduction in somatic symptoms from baseline across all three time points with a mean decrease of −8.5 (95% CI: − 11.4, −5.7) at 4 weeks, −9.4 (95% CI: −11.9, −6.9) at 8 weeks, and −10.9 (95% CI: −13.5, −8.3) at 13 weeks (all p<0.001; see Tables 2, 3; Figure 2; Supplemental Figure 1). The largest percent change from baseline was seen at 13 weeks, with a median percent decrease of 55% (IQR: 29%, 76%); median percent decrease from baseline at 4 and 8 weeks was 46% for both (IQR for 4 weeks: 16%, 63%; for 8 weeks: 21%, 65%). All participants had some numeric improvement from baseline to week 13. The individual components of the SSS-8 from baseline to week 13 are displayed in Supplementary Table 2. We also found similar improvements when looking only at the GI component of the SSS-8 with a mean decrease of −0.6 (95% CI: −1.1, −0.1), at 4 weeks, − 1.0 (95% CI: −1.5, −0.4) at 8 weeks, and −0.9 (95% CI: −1.6, −0.2) at 13 weeks, all p<0.015.

**Table 2.** Mean and Median Differences in Outcomes.

| Variable | 0-4 weeks | 0-8 weeks | 0-13 weeks |
| --- | --- | --- | --- |
| SSS-8 (mean difference compared to baseline) | -8.5 (95% CI: -11.4, -5.7), $p<0.001$ | -9.4 (95% CI: -11.9, -6.9), $p<0.001$ | -10.9 (95% CI: -13.5, -8.3), $p<0.001$ |
| FSS-9 (median difference compared to baseline) | -21 (95% CI: -26, -12), $p<0.001$ | -18 (95% CI: -24, -11), $p<0.001$ | -20 (95% CI: -28, -12), $p<0.001$ |
| Dyspnea (median difference compared to baseline) | -17 (95% CI: -35, -6), $p<0.001$ | -21 (95% CI: -33, -16), $p<0.001$ | -27 (95% CI: -38, -17), $p<0.001$ |
| Average pain (mean difference compared to baseline) | -1.7 (95% CI: -2.6, -0.7), $p=0.002$ | -1.6 (95% CI: -2.8, -0.4), $p=0.010$ | -2.2 (95% CI: -3.3, -1.0), $p<0.001$ |
| Pain intensity (mean difference compared to baseline) | -1.8 (95% CI: -2.7, -0.9), $p<0.001$ | -2 (95% CI: -3.1, -0.9), $p=0.001$ | -2.4 (95% CI: -3.4, -1.3), $p<0.001$ |
| Pain interference (median difference compared to baseline)<sup>a</sup> | -20.5 (95% CI: -31, -7), $p<0.001$ | -21.5 (95% CI: -37, -6), $p<0.001$ | -23 (95% CI: -35, -16), $p<0.001$ |
| PASS-20 (mean difference compared to baseline) | -16.4 (95% CI: -25.2, -7.6), $p<0.001$ | -21.1 (95% CI: -31.4, -10.9), $p<0.001$ | -28.6 (95% CI: -37.7, -19.5), $p<0.001$ |
| PROMIS-29 (median difference compared to baseline) | -4 (95% CI: -6, -3), $p<0.001$ | -5 (95% CI: -7, -3), $p<0.001$ | -4 (95% CI: -7, -3), $p<0.001$ |
| Brain fog (median difference compared to baseline) | -1 (95% CI: -2, 0), $p<0.001$ | -1 (95% CI: -2, 0), $p=0.002$ | -1 (95% CI: -2, -1), $p<0.001$ |
| SSS-8 GI (mean difference compared to baseline) | -0.6 (95% CI: -1.1, -0.1), $p=0.013$ | -1.0 (95% CI: -1.5, -0.4), $p=0.001$ | -0.9 (95% CI: -1.6, -0.2), $p=0.009$ |
| *FSS-9 visual analog scale (VAS; mean difference compared to baseline)* | *32.5 (95% CI: 21.1-43.9), $p<0.001$* | *26.8 (95% CI: 14.8-38.9), $p<0.001$* | *33.5 (95% CI: 23.0-44.1), $p<0.001$* |
| *\*increase represents better health* |  |  |  |

<sup>a</sup>1/23 (4%) missing values at 4 and 8 weeks

**Table 3.** Outcome Scores over Time.

| **Variable** | **Week 0** | **Week 4** | **Week 8** | **Week 13** |
| --- | --- | --- | --- | --- |
| SSS-8 (mean±SD) | 20.2±4.8 | 11.7±5.9 | 10.8±5.8 | 9.3±5.9 |
| FSS-9 (median, IQR) | 52 (44, 55.5) | 28 (20, 37) | 32 (26.5, 36.5) | 29 (12.5, 37.5) |
| Dyspnea (median, IQR) | 54 (26.5, 60.5) | 19 (6.5, 26) | 15 (4, 36) | 11 (1, 30.5) |
| Average pain (mean±SD) | 5.4±1.5 | 3.8±2.1 | 3.8±2.3 | 3.3±2.2 |
| Pain intensity (mean±SD) | 5.1±1.4 | 3.3±2.1 | 3.1±1.9 | 2.7±2.0) |
| Pain interference (median, IQR) | 43 (33, 52) | 20.5 (6, 31) | 14.5 (2, 34) | 10 (0, 28) |
| PASS-20 (mean±SD) | 44±17.9 | 27.6±16.1 | 22.8±15.4 | 15.3±14.4 |
| PROMIS (median, IQR) | 13 (8, 15) | 8 (5, 9) | 6 (5, 9) | 6 (4, 9) |
| Brain fog (median, IQR) | 3 “quite a bit”<br>(3, 4) | 2 “somewhat”<br>(1, 3) | 2 “somewhat”<br>(0.5, 3) | 1 “a little bit”<br>(1, 2) |
| SSS-8 GI (mean±SD) | 2.0±1.3 | 1.4±1.4 | 1.0±1.1 | 1.1±1.2 |
| FSS-9 VAS (mean±SD) | 27.3±13.8 | 59.8±23.6 | 54.1±24.0 | 60.8±23.4 |

**Figure 2.**
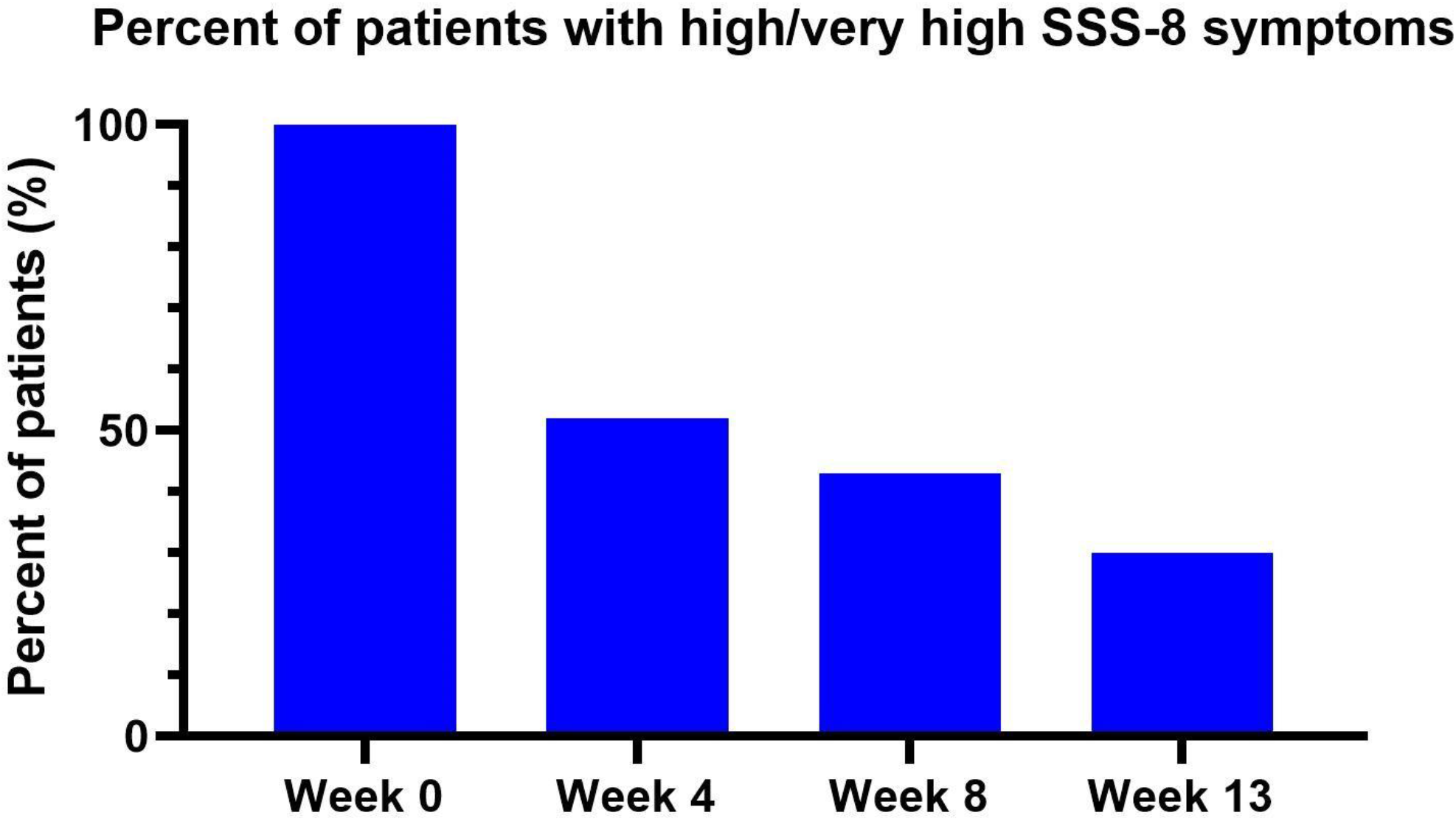
Percent of Participants with High/Very High SSS-8 Symptoms. Scoring of the SSS-8 is categorized as follows: no to minimal (0-3 points), low (4-7 points), medium (8-11 points), high (12-15 points), and very high (16-32 points) somatic symptom burden.

### Secondary Outcomes

For the other key secondary endpoints, we found a statistically significant reduction in symptoms from baseline in terms of fatigue, dyspnea, pain, pain anxiety, physical functioning, and brain fog across all three time points (Tables 2 and 3).

With respect to the question of whether exercise induced fatigue (as asked by the FSS-9), participants had a decrease in this belief at all three time points;this reduction was statistically significant at weeks 4 and 13 (median difference from baseline at 4 weeks: −2 (95% CI: CI: −3, − 1), p<0.001; at 8 weeks: −2 (95% CI: −3, −1), p=0.085; at 13 weeks: −2 (95% CI: −3, −1), p=0.010).

At baseline, thirteen (57%) participants indicated that they strongly agreed with the FSS statement that exercise made their symptoms worse. By four weeks, only two (9%) participants strongly agreed. This number continued to drop with only one (4%) participant strongly agreeing with the statement at eight and thirteen weeks.

When asked to visualize scenarios in which they often felt symptoms, the majority of participants had symptoms occur from imagining the activity (Supplemental Table 3).

Two participants suffered substantial weight loss (about 20-30 pounds) prior to entry into the study secondary to food intolerances with one requiring a nasogastric feeding tube for approximately 3 months. Both had an extensive medical evaluation which did not reveal an organic cause of weight loss. By the end of the intervention period, the food intolerances for both of these participants resolved, the feeding tube was not needed for the one participant, and weight was regained.

Two participants developed acute COVID-19 while participating in the program. Despite this, neither showed worsening of symptoms by week 13 (and instead showed numerical improvement).

## Discussion

The purpose of this study was to examine the efficacy of PSRT in reducing somatic symptom burden and other secondary outcomes such chronic fatigue, functional activity, average pain, pain intensity, anxiety from pain, “brain fog,” and dyspnea, in patients diagnosed with PASC Syndrome. We found that PSRT provided a statistically significant and clinically meaningful reduction (36) in somatic symptoms within 4 weeks and persisted through 8 and 13 week measurements. In addition, PSRT statistically significantly reduced fatigue, dyspnea, average and intensity of pain, pain anxiety, pain interference with activities, and improved activity levels.

In our study, each participant served as their own control; thus, there was no direct comparison group. However, the median duration of symptoms prior to entering the study was 267 (IQR: 144, 460) days, participants had seen a median of 3 (IQR: 2, 3) physicians prior to entry into the study, and they had tried a large variety of different therapies (Supplemental Table 3), including 52% having tried a mind-body therapy. These data suggest that the cohort represented a group of people with refractory PASC who would be unlikely to have a marked reduction in symptoms in a short period of time without intervention. In addition to the reported cohort-based findings, the SSS-8 score did not worsen in any participant from baseline to the completion of the program at 13 weeks.

At least one previous study showed that exercise may be helpful for PASC (37); however, this finding conflicts with anecdotal reports from patients and a recent report suggesting that exercise can worsen symptoms (37,38). In the current study, activity was encouraged beginning in the second week after the psychophysiologic education had been introduced. We hypothesize that the association of symptomatology with activity may have developed, in part, through a classical conditioning-like model. As such, ongoing activity without knowledge of the underlying process strengthened the conditioning and worsened or did not allow for improvement of symptoms. Our approach provided a method to break this cycle and desensitize participants from the conditioning (see desensitization portion of protocol in Supplementary materials). If this hypothesis is correct, it would explain why exercise worsens symptoms in some patients (and in previous studies) but was helpful in the current study. In the current study, participants were asked whether exercise worsened their symptoms. At baseline, the majority (57%) strongly agreed that exercise would worsen symptoms; however, only 4% of participants strongly endorsed this statement by the end of the study. While exercise-induced symptoms decreased, activity levels (as measured by PROMIS-29) increased. Therefore, while exercise increased during the treatment, participants’ symptoms and beliefs about the relationship between exercise and symptom-onset decreased.

During the program, we utilized visualization techniques to help desensitize participants to symptomatology when encountering triggering stimuli. Specifically, participants would visualize a physical activity that would typically bring on symptoms such as walking across the room or up the staircase. This visualization brought on symptoms in the majority of cases (Supplemental Table 2). This technique served a dual role of helping participants recognize that symptomatology could be generated by their mind but also served as an exposure mechanism to break the links between exertion and activity (see Supplementary materials for details). The finding that the majority of participants could reproduce symptomatology by visualization alone (i.e., without the actual activity) supports the concept of an underlying psychological component.

There have been a number of competing hypotheses about the underlying cause of PASC in patients without overt organ injury such as injury patterns not picked up through typical testing means (e.g., chest radiographs, computed tomography, MRI, skin biopsy). The injury patterns proposed include the development of microclots (11), mitochondrial dysfunction (39), or differences in capillary blood flow (40). In addition, a number of studies have reported immune system biomarker differences between Long COVID and controls. For example, one recent exploratory analysis reported differences in the immune phenotype in Long COVID patients (13). At present, these studies have yet to be confirmed; however, if a future study did validate one (or more) of these findings, this does not necessarily negate our psychophysiological hypothesis. For example, the emotion of embarrassment results in changes in capillary blood flow (i.e., microcirculation) and results in changes in the color of the skin (ie., flushing). This same pattern could hypothetically occur from chronic underlying emotions and in other tissue beds. Likewise, someone who is immobile for psychophysiologic reasons may be more likely to develop a microclot. Psychophysiologic syndromes also have been associated with changes to immune cells or predisposition to infection. One finding that would argue against our hypothesis would be the presence of chronically actively replicating virus with associated inflammatory changes. To date, that has not been identified, though some have reported the presence of a persistent spike protein. Of note, during the program, two participants developed acute COVID-19 but neither had worsening symptoms and both showed numerical improvement by the end of the end of the 13 week period.

The symptoms described in our participants and the PASC literature resemble previously reported reactions to traumatic experiences in patients without physical injury. For example, multiple reports dating back to the American Civil War era report physical symptoms from some soldiers returning from the battlefield with pain, dyspnea, fatigue, exertional fatigue, tachycardia, and sleep problems even when not wounded physically (21–23,41). In addition, PTSD has been associated with changes in cognition (42–44) and even changes in the brain as noted in MRIs (42,43). The existence of such psychophysiologic syndromes with a constellation of symptoms similar to PASC provides a historical precedent for a relationship between psychological states and these physical manifestations.

Although the CDC does not require proof of prior COVID-19 infection as part of their definition for PASC; however, having a population that definitively had COVID-19 was important to limit variability in our sample. With that context, the most common reason for exclusion from the study was the lack of a positive test for COVID-19 (see Figure 1). However, the presence of identifiable organ injury from acute COVID was one of the least common reasons for exclusion. This pattern of exclusions is consistent with our overall scientific premise. Of note, the constellation of symptoms described in this report can also represent serious pathology; due to this, all participants were evaluated by a physician (s) prior to screening by the study team, which also included a physician.

### Limitations and Future Directions

The limitations of the current study include the small sample size and the lack of a comparator group. The later concern is mitigated somewhat for the reasons noted above and particularly in that these participants seem to have been refractory in their symptoms. During participant recruitment, the study was advertised as a mind-body intervention resulting in people who are open to this type of intervention self-selecting to participate, which limits generalizability due to selection bias. Other limitations include the subjective reporting of symptoms, which we attempted to mitigate by using validated tools measuring multiple domains. The optimal duration and content of the program remains unknown, including the possibility that there could be differential needs depending on individual participants. The course was taught by the same two instructors with both generally present during the first 5 weeks and one teaching the MBSR portion during the final 9 weeks. Therefore, we cannot assess the effect the intervention may have had if administered by different instructors. The measure of brain fog used has not been externally validated but we felt was important to capture since it was reported as a common symptom in the PASC literature. Future work may consider the incorporation of this parameter into the SSS-8 questionnaire and validating the overall tool. Despite the efficacy of PSRT seen in this study, our results do not necessarily prove the scientific premise of the psychophysiologic model. Specifically, PSRT could theoretically be helpful even if there was an underlying primary organic source of symptoms. However, the techniques utilized and described rely heavily on this psychophysiologic premise. Lastly, the population studied focused on patients without identified organ injury and should not be generalized to other populations where organ injury is present (e.g., lung fibrosis, myocarditis, cerebrovascular accident, demyelination disease, or any of a number of other processes).

## Conclusions

In this cohort of PASC patients without identified organ injury, PSRT reduced symptomatology across multiple domains within four weeks; this benefit persisted over the 3 month study period.

## Data Availability

All data produced in the present study that are able to be appropriately de-identified are available upon reasonable request to the authors.

## Acknowledgements

This study was funded via philanthropic support from Adam D’Angelo and James O’Shaughnessy. We would like to also thank Antoine Dusséaux, Sarah Rainwater and Deborah Bliss as contributors to the Psychophysiologic Research Group.

