## Supplemental Materials for "Psychophysiologic symptom relief therapy (PSRT) for post-acute sequelae of COVID-19: a non-randomized interventional study"

**Supplemental Table 1. PASC treatments utilized prior to study start**

| Medications | Physical | Other |
|---|---|---|
| Bupropion<br>Duloxetine<br>Budesonide-formoterol<br>Gabapentin<br>Pregabalin<br>Nortriptyline<br>Topiramate<br>Colchicine<br>Diclofenac<br>Cortisone injections<br>Fremanezumab<br>Maraviroc<br>Acyclovir<br>Pravastatin<br>Prednisone<br>Methylphenidate<br>Pyridostigmine<br>Benzodiazepines<br>Beta-blockers<br>Antihistamines<br>Antibiotics | Massage therapy<br>Dry needling<br>Physical therapy<br>Acupuncture<br>Positional release therapy<br>Cryotherapy | Supplements<br>Mindfulness course<br>Therapy (general psychotherapist) |

**Supplemental Table 2. Median values of components of SSS-8 over time**

| **Component*** | **Baseline** | **Week 4** | **Week 8** | **Week 13** |
|---|---|---|---|---|
| Gastrointestinal (median, IQR) | 2 (1, 3) | 1 (0, 2) | 1 (0, 2) | 1 (0, 2) |
| Back (median, IQR) | 2 (1, 3) | 1 (1, 3) | 2 (0, 2) | 1 (0, 3) |
| Joint pain (median, IQR) | 3 (1, 3) | 1 (0, 3) | 1 (0, 2) | 0 (0, 1) |
| Headache (median, IQR) | 3 (2, 4) | 1 (1, 2) | 1 (0, 2) | 1 (0, 2) |
| SOB (median, IQR) | 2 (1, 3) | 1 (1, 2) | 1 (0, 1) | 0 (0, 2) |
| Dizzy (median, IQR) | 2 (1, 3) | 1 (0, 1) | 0 (0, 1) | 0 (0, 1) |
| Tired (median, IQR) | 4 (3, 4) | 1 (1, 3) | 2 (1, 3) | 1 (0, 3) |
| Sleep (median, IQR) | 3 (2, 4) | 2 (0, 3) | 1 (0, 3) | 2 (1, 3) |

*For each SSS-8 component, 0 represents that issues are "not at all" present, 1 that they are "a little bit" present, 2 that they are "somewhat" present, 3 that they are "quite a bit" present, and 4 that they are "very much" present.

**Supplemental Table 3. Presence of participant symptom(s) during visualization exercise**

| **Participant** | **Reviewer 1** | **Reviewer 2** |
|---|---|---|
| A | No | Unclear/uncertain |
| B | Unclear/uncertain | Unclear/uncertain |
| C | Unclear/uncertain | Unclear/uncertain |
| D | Unclear/uncertain | Yes |
| E | Unclear/uncertain | Yes |
| F | Yes | Unclear/uncertain |
| G | Yes | Yes |
| H | Yes | Yes |
| I | Yes | Yes |
| J | Yes | Yes |
| K | Yes | Yes |
| L | Yes | Yes |
| M | Yes | Yes |
| N | Yes | Yes |
| O | Yes | Yes |
| P | Yes | Yes |
| Q | Yes | Yes |
| R | Yes | Yes |
| S | Yes | Yes |
| T | Yes | Yes |
| U | Yes | Yes |
| V | Yes | Yes |
| W | Yes | Yes |

Participant symptom visualization refers to a desensitization technique where participants are asked to visualize a movement or action that typically induces symptoms. Two independent reviewers (M.W.D. and P.H.) retrospectively evaluated whether the participant was able to induce symptoms.

**Supplemental Figure 1. Median scores in outcome measures over time with percent reduction between baseline and 13 weeks**

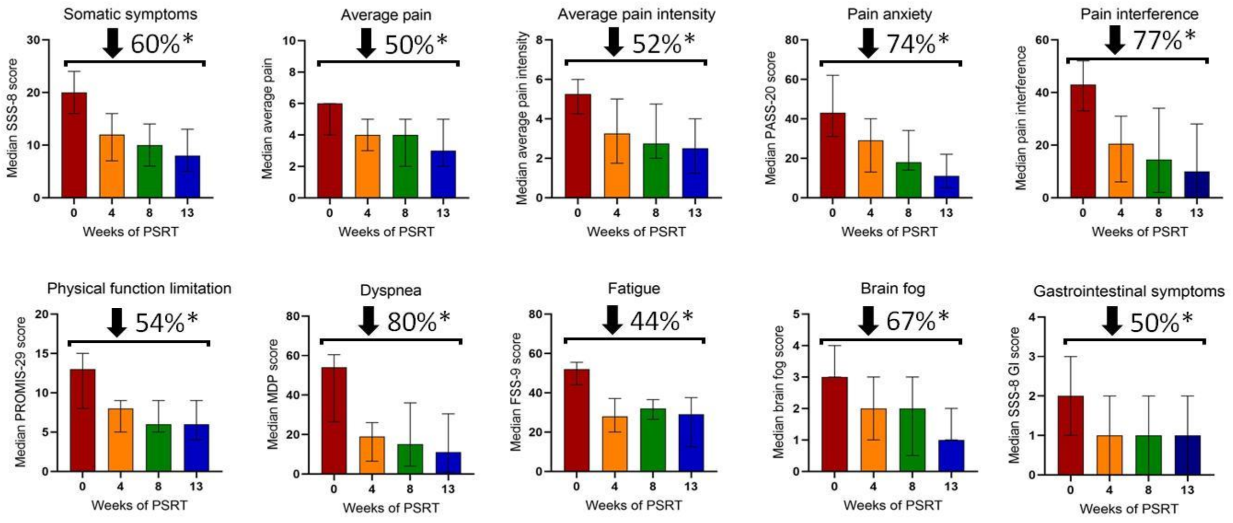


Percent reduction represents the reduction between the overall group median at baseline and the overall group median at 13 weeks. *represents a p-value of <0.015 from an unpaired analysis using Wilcoxon rank-sum test comparing group medians at baseline to group medians at week 13

**Psychophysiologic Symptom Relief Therapy (PSRT) Treatment Protocol**

PSRT is based on the notion that nonspecific pain and idiopathic symptoms are a result of psychophysiological processes. The PSRT intervention consists of a single one-on-one session with an instructor, group educational and skills-training sessions, and a mindfulness-based stress reduction (MBSR) program which includes one full-day (approximately 6 hours) session/retreat. The group educational and skills training sessions are held twice per week for four weeks with each session lasting 1.5-2 hours. The MBSR program consists of one 1.5-2 hours class per week for eight weeks (in addition to the full-day MBSR retreat and an orientation class). The PSRT treatment is delivered in small groups facilitated by trained instructors. Participants are provided with Worry Less Live More by Lizabeth Roemer and Susan Orsillo (1), The Mind Body Prescription by Dr. John Sarno (2), and Unlearn Your Pain by Dr. Howard Schubiner (3) to read as part of material for the course. The educational and skills training sessions are focused on understanding the relationship between psychological stress and pain or other symptoms and are divided in four components according to their content:

**Component I: Psychophysiologic Education**

The goal of this component is to help participants recognize that their symptoms are part of a psychophysiologic process instead of arising solely from a physical etiology. Recognition of the psychophysiologic process is accomplished through education on the relationship between the mind and body, exploration of participants' pain and symptom history, and identifying “inconsistencies” in the experience of symptoms or pain (such as variation in symptom location or triggers). Understanding the relationship between psychological stressors and symptomatology is emphasized throughout the intervention. By identifying patterns of increased stress exacerbating symptoms, participants are able to appreciate the contribution of underlying stressors to their conditions. For example, one participant had pain when walking upstairs while at work but recognized (upon reflection during the course) that he did not have pain when walking up even more stairs when on a vacation. Through these experiences, participants understand the connection between psychological processes. In addition, the participants explore their own stressors, some of which they recognized consciously and others only recognized as part of reflection during the program.

*Examples:*
In the above example, a participant recognized an inconsistency in their experience of pain. When walking up steps at work he had pain but when he was walking up steps while on vacation, he felt no pain. This realization was an epithetical moment for the participant during the intervention. They started to re-incorporate more activity and be more engaged in developing psychological techniques aimed at reducing their psychophysiologic pain.

Another participant experienced a multitude of symptoms while in the workplace but felt far fewer when at home. When this participant recognized that the onset of their symptoms was environmental and, in turn, driven by workplace stress, they better understood that their symptoms were psychophysiological instead of arising from a physical abnormality.

**Component II: Desensitization (including visualization) and returning to physical activity**

Conditioned responses of pain and/or symptoms, perpetuated by psychological underpinnings, can arise after physical triggers and remain after the initial trigger subsides. Similar to the classical conditioning model, a neutral stimulus can become a symptom-inducing trigger when coupled with a pain/symptom-inducing stimulus. Therefore, a neutral stimuli can become associated with pain or other symptoms without nociceptive input. For example, muscle tension previously arising from pain may become associated with neutral stimuli like sitting or walking leading to fear, avoidance behavior, and restriction. As a result, a key portion of our intervention is "desensitization"; these techniques are aimed towards breaking the cycle of pain/symptoms and decoupling the fear of symptoms with the neutral stimuli.

Visual motor imagery (visualization of a symptom-inducing situation) is a desensitization technique where participants are asked to visualize a movement or action that typically induces symptoms. This visualization often brings on symptoms. When visualization induces symptoms, the notion that symptoms or pain are a result of a psychophysiologic process is reinforced. Participants are encouraged to repeatedly visualize movements or actions, without physically moving, while also engaging in self-soothing behavior like affirmational statements. This repetitive exposure to visualization-induced symptoms ultimately reduces symptoms until participants are no longer able to experience symptoms through visualization. At this point, when symptoms are no longer evoked from visualization, participants may begin to incorporate the movements or actions that they had visualized as inducing symptoms (example below).

Another crucial component of desensitization is identifying the movements, actions or environments in a participant's daily life that have been conditioned to induce symptoms or pain. For example, if sitting triggered symptoms, participants would sit and repeat the knowledge that their symptoms arising from sitting is conditioned instead of a response to a physical issue. With repeated exposure and practice, the neutral stimuli and pain/symptom response are decoupled or deconditioned and symptoms subside. Participants are then able to incorporate tasks and activities that they previously avoided. Through a successful return to daily life and activities, the "knowledge therapy" component of PSRT is reinforced as participants recognize that their symptoms do not arise from physical triggers but psychological ones. Participants can then safely return to activities under the supervision of a physician (example below).

*Examples:*
When instructed to imagine walking across the room, while still sitting in their chair, multiple participants developed symptoms including shortness of breath, chest pain and the onset of a headache. This experience allowed the participants to recognize the psychological component of their symptoms which served both to strengthen the knowledge therapy portion of the program but also provided a means for beginning the desensitization (or decoupling) of the symptoms from the triggers. From this, participants were more committed to the intervention and could use visual imagery coupled with self-affirming techniques to decouple the symptomatology from the trigger. When successfully decoupled through this visualization, participants were able to walk across the room with fewer symptoms and were able to continue the desensitization process.

One participant was having pain in their hand and wrist while journaling about a stressful situation. The journal exercises were then intentionally changed to be about a joyful experience. Upon completing the exercise, the participant recognized that the pain did not come on while writing about a joyful experience. The sequence was repeated with the same result. After the participant recognized this relationship, the writing toward stressors was continued but with self-affirmation and recognition of the true causal reason for the pain. Over time, the association with writing was broken.

**Component III: Emotional expression - psychology of the syndrome**

This emotional expression component of the treatment occurs in conjunction with the education and desensitization components. In 1959, the idea that chronic pain or chronic physical symptoms may be precipitated and perpetuated by avoided emotions and negative thought processes (e.g., anger that participants do not acknowledge or address) was described (4). Such ideas about emotions and physical symptoms have been supported by recent research (5–10). Treating a psychophysiologic disorder requires appreciation of factors that exacerbate chronic symptoms such as conflict and emotional avoidance, as well as incorporation of emotional expression strategies. Activities aimed at improving emotional expression, like writing exercises, journaling and self-reflection, give participants an opportunity to express avoided emotions which have previously exacerbated symptoms (11). Emotional Awareness and Expression Therapy (EAET), developed and tested by Lumley, Schubiner, and colleagues (6,12–15), includes emotional expression concepts similar to those utilized in PSRT. The overall goal of emotional expression exercises in PSRT is to help participants experience and express a wide range of emotions, including anger, sadness, and positive emotions such as joy. Emotional expression exercises are introduced to participants during the psychophysiologic education component of the treatment and continue to be utilized until participants enter the final component of PSRT (mindfulness; described below). For example, participants are shown free writing exercises and encouraged to write about a stressor of their choice or write about a specific emotion (e.g., joy). Participants are also introduced to writing dialogues (writing as if they are two people interacting about a specific situation), and to writing an "unsent letter" where they write an expressive letter to a significant person (without actually sending the letter). Participants complete these emotional expression exercises at various points during the sessions and are also encouraged to write outside of the treatment sessions. Participants can discuss their reactions to the process of completing the writing exercises during sessions.

**Component IV: Stress reduction - Mindfulness Based Stress Reduction (MBSR)**

The last nine weeks of the intervention are oriented towards developing techniques for stress reduction while continuing to practice what has been learned in the earlier weeks of the treatment. The knowledge gained in the earlier weeks along with the improvement of activities and symptoms allows for the optimal environment for this portion of the program. Mindfulness Based Stress Reduction (MBSR) is the final portion of the intervention; MBSR was created at the Center for Mindfulness at the University of Massachusetts Medical Center by Jon Kabat-Zinn in an effort to prevent and treat stress-related conditions (16). Studies have shown that MBSR results in improved health and wellbeing and has been effective in reducing stress, anxiety, depression, and chronic pain (11,17–19). The MBSR program, which adheres to the protocol as outlined by the Center for Mindfulness at the University of Massachusetts (20), includes a weekly 1.5-2 hour class, mindfulness practices, and encouraged daily home practice. During weekly classes, participants learn strategies aimed at ‘present moment awareness’ (11) so that they may acknowledge and practice acceptance of what they are experiencing in the present moment. Present moment awareness encourages participants to observe their thoughts and prevent emotional reactivity; this leads to balanced emotions, a sense of calm, and improved wellbeing. Some of the specific strategies included in the classes are education about stress reactivity, learning to respond to stressors, practicing awareness of breath, body scan, Hatha yoga, and sitting meditation.
